# Micro-costing Diagnostics in Oncology: From Single-Gene Testing to Whole Genome Sequencing

**DOI:** 10.1101/19009969

**Authors:** Clémence TB Pasmans, Bastiaan BJ Tops, Elisabeth MP Steeghs, Veerle MH Coupé, Katrien Grünberg, Eiko K de Jong, Ed MD Schuuring, Stefan M Willems, Marjolijn JL Ligtenberg, Valesca P. Retèl, Hans van Snellenberg, Ewart de Bruijn, Edwin Cuppen, Geert WJ Frederix

## Abstract

**Purpose:** Predictive diagnostics play an increasingly important role in personalized medicine for cancer treatment. Whole genome sequencing (WGS) based treatment selection is expected to rapidly increase worldwide. Detailed and comparative cost analyses of diagnostic techniques are an essential element in decision-making. This study aimed to calculate and compare the total cost of currently used diagnostic techniques and of WGS in treatment of non-small cell lung carcinoma (NSCLC), melanoma, colorectal cancer (CRC) and gastrointestinal stromal tumor (GIST) in the Netherlands.

**Methods:** The activity-based costing (ABC) method was conducted to calculate the total cost of included diagnostic techniques based on data provided by Dutch pathology laboratories and the Dutch centralized cancer WGS facility. Costs were allocated to four categories: capital costs, maintenance costs, software costs and operational costs. Outcome measures were total cost per cancer patient per included technique, and the total cost per cancer patient per most commonly applied technique (combination) for each cancer type.

**Results:** The total cost per cancer patient per technique varied from € 58 (Sanger sequencing, 3 amplicons) to € 4738 (paired tumor-normal WGS). The operational costs accounted for the vast majority over 90 % of the total per cancer patient technique costs. The most important operational cost drivers were consumables followed by personnel (for sample preparation and primary data analysis).

**Conclusion:** This study outlined in detail all costing aspects and cost prices of current and new diagnostic modalities used in treatment of NSCLC, melanoma, CRC and GIST in the Netherlands. Detailed cost differences and value comparisons between these diagnostic techniques enable future economic evaluations to support decision-making on implementation of WGS and other diagnostic modalities in routine clinical practice.

## Introduction

Newly developed medicines (targeted therapies and immunotherapies) play an increasingly important role in treatment of cancer (1–2). However, only subgroups of patients respond to these (mostly expensive) treatments (3–6). Patients who do not respond can experience serious side effects. Matching each patient to the appropriate therapy is complex and as a consequence, not all patients receive the treatment they could have benefitted from (3–5). This calls for a better patient selection, improvement of personalized treatment and thereby expectantly improving the patients’ life expectancy, experienced quality of life and reducing health care costs. Optimal predictive diagnostics in molecular pathology are necessary to determine which therapy is most appropriate for a patient (7–10).

In predictive diagnostics of somatic molecular analyses in pathology, various techniques can be used to depict genetic characteristics of a tumor. Single-gene analysis or sequencing of targeted gene panels (TGP) using next generation sequencing (NGS) techniques, or a combination of the two, are routine practice in the diagnostics trajectory for different cancer types (11). In current clinical practice, there is large variation in both the frequency and type of technique used for the selection of cancer treatment (12). In comparison, the advanced diagnostic technique of whole genome sequencing (WGS) is currently only applied in research context in oncology.

The main advantage of WGS, in contrast to TGP, is that it is able to detect all types of DNA alterations (i.e. mutations, copy number alterations, structural variants, tumor mutational burden and DNA repair status) of the tumor (13–14). It increases the chance of optimal treatment selection, and determines eligibility of patients for clinical trials as many study inclusion markers are not included in standard diagnostic gene panels. From a technical point of view, WGS could therefore replace a multitude of currently used diagnostic techniques, but it comes at a higher cost. Recent costing studies have indicated that costs range from € 265 to € 309 for single-gene techniques (12), from € 376 to € 968 for small TGPs (5–50 gene panels (15–16), ∼ 50 gene panels (12)), and from € 333 to € 1948 for larger TGPs (> 50 gene panels (15), 90 gene panels (17)) in 2014–2015. The cost of WGS per cancer patient (paired tumor-normal) was estimated at € 6676 (17) and at € 5645 (18) in 2015–2016, at $ 4484 (about € 3870) in 2017 (19), and at £ 6841 (about € 7501) in 2019 (20) (€ 1669,€ 1411, $ 1121, £ 3420 per genome equivalent, respectively).

These studies performed micro-costing analyses. Some studies specifically made use of the activity-based costing (ABC) method, a process-based cost allocation technique (21). Nevertheless, results are difficult to compare, as included cost components differed among all studies as well as the interpretation of process steps, and, as such, the incorporated related costs. Cost drivers are anticipated to be platform utilization (12, 18) and consumables (14, 15–18).

The implementation and use of WGS is expected to rapidly increase worldwide in the coming years (13–14). A variety of reasons underlie this prospect. Namely, the increasing availability of targeted drugs (1–2), and the increased registration of pan-cancer drugs (22), leading to an increase in needed tests and accumulation of sequential tests. Therefore, it is essential to determine both costs and effects of WGS-based treatment selection versus current practice in an economic evaluation. Such evaluations encompass complete overviews of all costs and effects involved in a specific disease area, from diagnostics to treatment and hospitalization. Detailed and comparative cost estimations of diagnostic techniques are required. These are important in assessing the added value (in monetary terms) of new diagnostic modalities, and in providing insight when to replace standard diagnostic techniques with these new modalities.

To the best of our knowledge, no previous research investigated the costs of currently used diagnostic techniques and WGS in the context of predictive testing in cancer treatment selection using a consistent and uniform costing method. Therefore, we performed a micro-costing study using similar process-based cost calculations of the different diagnostic techniques application across Dutch pathology laboratories (hereinafter referred to as labs) and WGS used in a central lab. We aimed to calculate and compare the total cost of currently used diagnostic techniques and of WGS in treatment of NSCLC, melanoma, colorectal cancer (CRC) and gastrointestinal stromal tumor (GIST) in the Netherlands.

## Methods

### Data availability

The data used for the study were obtained from 24 Dutch labs and the cancer WGS facility of Hartwig Medical Foundation (HMF) in the year 2018. The predictive diagnostic techniques included from the participating labs were techniques that are currently used for treatment selection of advanced NSCLC, melanoma, CRC or GIST. This led to the inclusion of following techniques: immunohistochemistry (IHC), Fluorescence In Situ Hybridization (FISH), pyrosequencing (Pyro seq), High Resolution Melting (HRM), Sanger sequencing (Sanger), NGS gene panels, Cobas and Biocartis. In this study, the costs of certain techniques were subdivided regarding their target genes (Sanger and Biocartis), cancer hotspot panels (NGS) or protein expression (IHC).

Included techniques were selected based on an inventory at participating labs. These labs received a questionnaire to obtain information about most frequently used techniques in treatment of the different cancer types. In addition, the frequency of technique usage was extracted from the nationwide network and registry of histo-and cytopathology in the Netherlands (PALGA) (23), which contains the digital pathology reports of all 46 Dutch pathology laboratories since 1971. The inventory was performed between 01-10-2017 and 30-09-2018 and was substantiated with feedback moments to the labs in order to ensure additional validity. Data on technique frequency usage are available in Supplementary Table 1.

Information about WGS was obtained from the HMF facility, a centralized independent organization focused on clinical-grade WGS of cancer patients. WGS-based analyses were performed as part of clinical trial studies involving treatment of all types of cancer patients, so regardless of cancer type.

### Micro-costing design

The cost calculations for the different diagnostic techniques used in the Netherlands were performed using the ABC method (21). For this purpose, a measurement plan was created including the essential cost components.

The most frequently used techniques in treatment of NSCLC, melanoma, CRC and GIST were defined by the participating 24 Dutch labs. Per technique, three labs, if possible, using the specific technique were consulted to determine the costs by filling out the measurement plan. In an organized meeting, consensus was reached between the labs concerning these measurement plans. Consensus was not based on averaging of cost prices of different labs, but rather based on an ‘average’ lab with realistic samples numbers, accepted protocols and equipment. The measurement plans were sent back and forth several times for feedback after this meeting. Additionally, supplier standard list prices were requested and received for consumables, and for acquisition and maintenance of the platforms. Together, this led to a cost overview with final cost prices related to the test, data analysis and reporting process per technique, per cancer type.

With regard to WGS, a measurement plan was completed by the HMF facility, which corresponded to the one filled out by the labs. The test, data analysis and reporting process was expressed in a final cost price. The final cost price estimation was based on utilization in a decentralized setting, or an average Dutch lab practice, and standard list prices of the supplier for acquisition and maintenance of the platforms and for consumables.

In sum, a so called standard case perspective was maintained in calculating the base case cost prices for all techniques for the purpose of realistic cost comparison. This means that an average lab practice was assumed, and suppliers’ standard list prices were used, concerning all techniques. The assumptions underlying the cost calculations are shown in Table 1, which are all based on the standard case perspective.

**Table 1.**
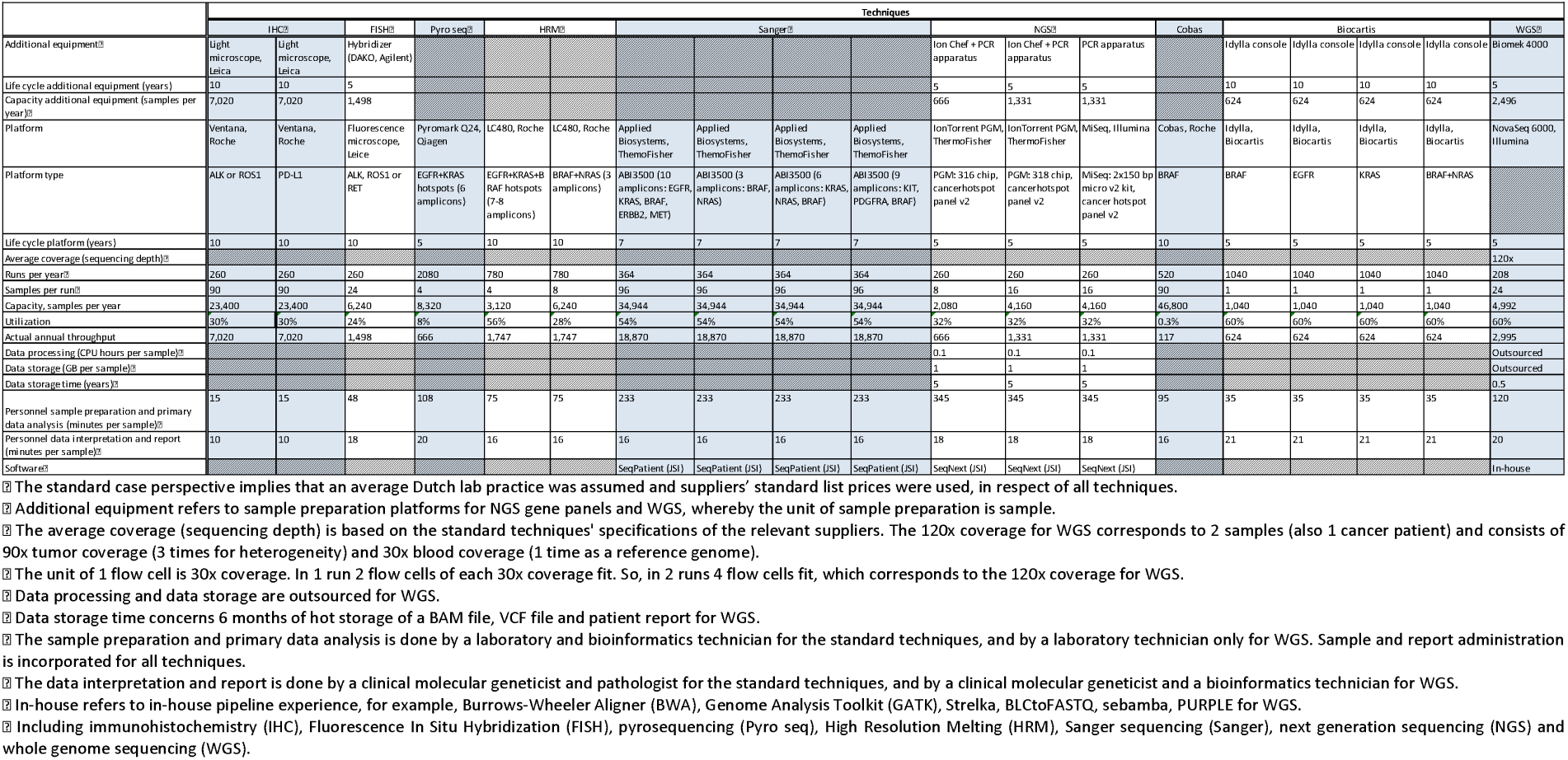
Base case assumptions for cost calculations of diagnostic applications based on the standard case perspective.⍰

### Allocation of costs

Costs directly related to the test, data analysis and reporting process were taken into account. Costs that are associated with obtaining the material (blood and tumor biopsy withdrawal), DNA extraction in both tumor and blood, and overhead costs were excluded. Included costs were allocated to four categories: capital costs, maintenance costs, software costs and operational costs.

Capital costs were fixed costs of the platforms. The life cycle, interest rate and annuity factor of the various platforms were used in calculating the annual capital costs per cancer patient. Maintenance costs were annual returning fixed costs for platform maintenance. No maintenance costs were taken into account for the first year as the platforms have a warranty for the first year. The annual maintenance costs were estimated per cancer patient for the other years. Software costs involved either software acquisition (license) costs or costs incurred for daily supervision and maintenance of the pipeline, and were calculated per cancer patient. Operational costs consisted of costs incurred for the process of analysis, such as consumables, personnel for sample preparation, primary data analysis, interpretation and report, and data processing and storage. These operational costs were also estimated per cancer patient. Finally, total cost per cancer patient was calculated by summing up all the calculated total costs per cost category.

### Analyses

#### Base case analysis

The base case analysis was performed from the earlier defined standard case perspective based on the assumptions described in Table 1. The primary outcome measure of interest was the total cost per cancer patient per included technique.

The second outcome measure included the total cost per cancer patient per most frequently applied technique (combination). Only those techniques used for targeted therapy stratification based on genomic aberrations were considered (so excluding IHC testing (programmed death ligand 1 (PD-L1) protein expression as is included in Table 1 and 2). The total cost per technique was calculated for each cancer type separately with a maximum of three different technique combinations ordered by technique frequency usage. WGS was included as a sole potential future practice for all cancer types for which combinations with other techniques were redundant.

**Table 2.**
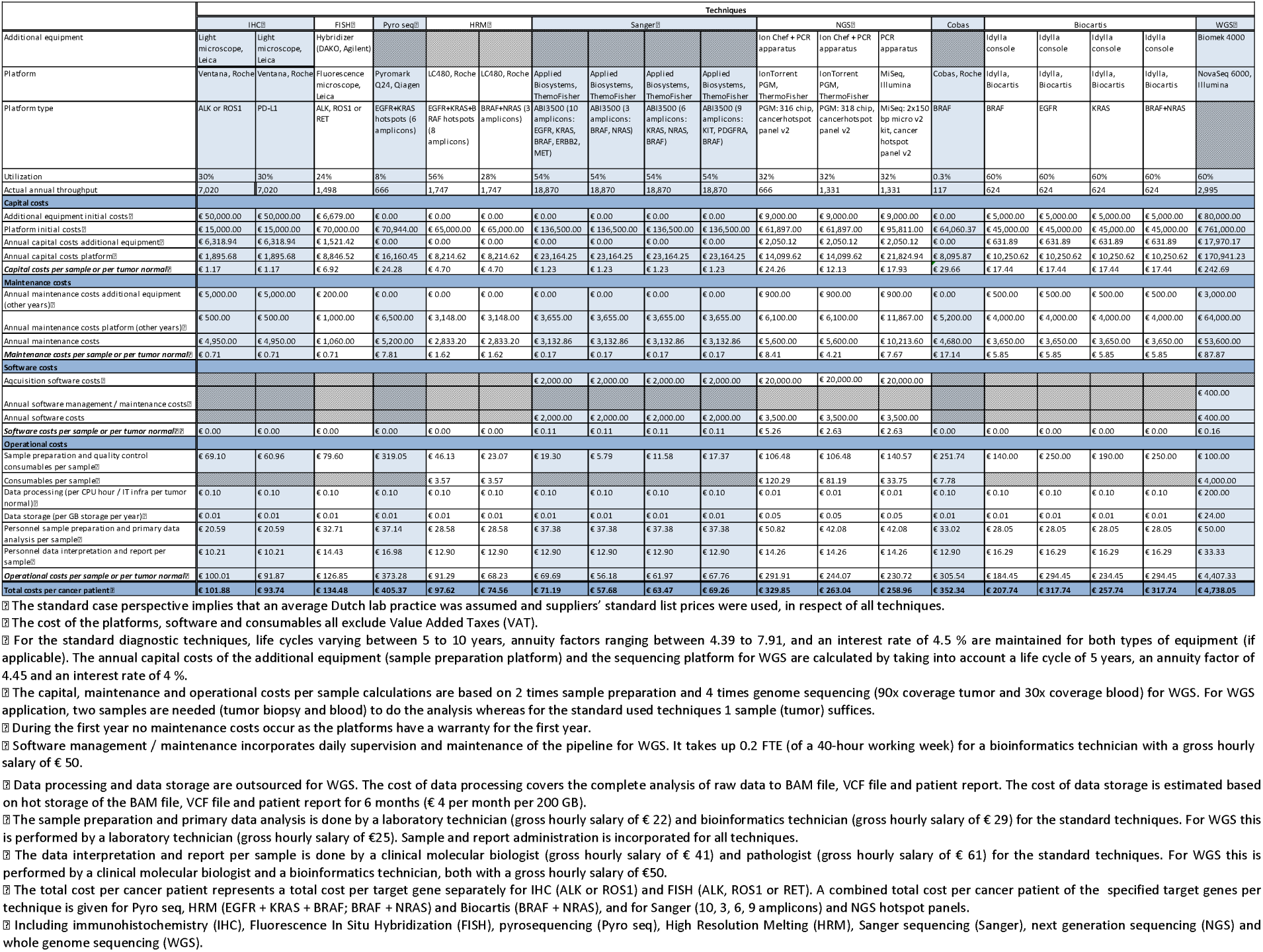
Process-based cost calculations of diagnostic applications based on the standard case perspective.⍰

For standard techniques to be included, the following condition had to be met: the technique should be performed in ≥ 2 labs (inventory labs) and included ≥ 5 % of the analyses in total for the respective cancer types (PALGA data; Supplementary Table 1). This led to the inclusion of NGS gene panels, Sanger, HRM, IHC and FISH. These techniques covered at least 80 % of the performed analyses per cancer type (Supplementary Table 1). In addition, WGS was included based on application at the HMF facility only.

#### Sensitivity analysis

In order to obtain a representation of the variation in technique usage and associated process costs, across all 24 labs, the distribution of costs were mapped around the average. Moreover, as the HMF facility is currently one of few WGS testing suppliers in the Netherlands, their actual practice was taken into account as a sensitivity analysis. Finally, two anticipated cost drivers were selected for this sensitivity analysis: utilization of the platforms and the cost of consumables, based on previous research (12, 14, 15–18). The extent of variation of these parameters were based on lab- and HMF-specific practices in the year 2018.

For the sensitivity analyses, only techniques included in the base case analysis concerning the second outcome measure were taken into account. A margin of + 15 % and – 15 % around the calculated average platform utilization for the standard techniques applied by different included labs was deemed to be a realistic variation. Therefore, utilization of the platforms varied from 17 to 47 % for NGS gene panels (average 32 %), 39 to 69 % for Sanger (average 54 %), 13 to 43 % for HRM (average 28 %), 15 to 45 % for IHC (average 30 %) and 9 to 39 % for FISH (average 24 %). For WGS, the average platform utilization was varied to the actual practice use by + 30 %: from 60 % to 90 %. The cost of consumables was reduced in the sensitivity analysis by 30 % for all the techniques, which was based on the reasonable expectation of discounts from the suppliers.

## Results

The assumptions underlying the cost calculations for the application of the included diagnostic techniques are depicted in Table 1. The Table shows all values of the various factors based on the standard case perspective as outlined for both standard diagnostics and WGS below.

### Standard diagnostics

For each of the included standard diagnostic techniques one tumor sample is needed, corresponding to the test of one cancer patient. The number of samples that can be analyzed per run and the sequencing depth were based on the concerning supplier specifications. Furthermore, suppliers’ standard list prices were used as cost of acquisition and maintenance of the platforms, and as cost of consumables. Utilization of the sequencing platform, personnel time needed for sample preparation, primary data analysis, data interpretation and report, are all based on the standard practice of an average lab using the technique. Gross hourly salaries of the laboratory technician, bioinformatics technician, clinical molecular biologist, and pathologist were based on Dutch hospital collective employment agreement 2018 costs.

### WGS

The calculations for WGS are based on the sequencing platform NovaSeq 6000 from Illumina, which is used in the HMF facility. Furthermore, a liquid handler is included for sample preparation. Per cancer patient two samples are needed for the sequence analysis: one tumor and one reference (blood) sample, which allows the necessary tumor to normal comparison. The sequencing unit for WGS is a sequencing depth of 30x coverage. In applying WGS, two sample preparations and four sequencing units are needed. The four sequencing units include three times 30x coverage of the tumor (to compensate for tumor purity heterogeneity) and one time 30x coverage of the reference sample. Acquisition and maintenance costs of the platforms are based on utilization of the technique in an average Dutch lab practice. The costs of consumables are based on Illumina’s standard list prices, not taking into account discounts. In line with the standard case perspective, the number of samples per run and runs per year are 24 and 208, respectively, with a 60 % utilization of the sequencing platform. Furthermore, personnel time needed for sample preparation, primary data analysis, data interpretation and report, are all based on standard practice of an average lab. Data processing and data storage are outsourced and concerns processing and storage of BAM files, VCF files and patient reports. Gross hourly salaries of the laboratory technician, clinical molecular biologist, and bioinformatics technician were based on the HMF facility employers’ 2018 costs.

Concerning all techniques, costs of acquisition and maintenance of the platforms, any software acquisition (license), and used consumables exclude Value Added Taxes (VAT) (Dutch standard rate is 21 %). Utilization percentages are defined based on 100 % utilization, indicating that the platforms run samples 8 hours a day and 5 days per week (average working week). All costs are reported in 2018 euros.

### Base case analysis: Primary outcome

Main cost components and outcomes are displayed in Table 2. A detailed cost overview including all measured cost items is available and enclosed as Supplementary Table 2. The total cost per cancer patient per technique varied from € 58 (Sanger, 3 amplicons) to € 4738 (paired tumor-normal WGS). For most techniques, total per cancer patient costs (including any target gene, hotspot panel or protein expression subdivisions) were over 90 % attributable to operational costs. Within this cost category, the most important cost drivers were consumables (for most > 50 % of operational cost) followed by personnel for sample preparation and primary data analysis.

### Base case analysis: Secondary outcome

Table 3 depicts a cost overview of most frequently occurring technique (combinations), including only those techniques used for targeted therapy stratification based on genomic aberrations, focusing on NSCLC, melanoma, CRC and GIST. The WGS technique would be a potential future (combinational) indication of practice use with a year 2018 total cost per cancer patient of € 4738. For WGS no additional IHC or FISH for detection of fusion genes (e.g. EML4-ALK) is necessary as is required for NGS gene panels. However, for immunotherapy, sequencing techniques like NGS and WGS would have to be applied in combination with IHC protein expression (PD-L1) testing of the tumor, which is not included in Table 3. For the specific cancer types, the total cost per cancer patient varied between € 58 (Sanger) and € 284 (NGS) for melanoma, € 63 (Sanger) and € 284 (NGS) for CRC, € 69 (Sanger) and € 284 (NGS) for GIST, and technique combinations for NSCLC ranged from € 313 (Sanger and FISH) to € 526 (NGS and FISH).

**Table 3.** Costs of frequently applied combinations of techniques per cancer type.⍰

|  |  |  |  |  |  |  |  |
| --- | --- | --- | --- | --- | --- | --- | --- |
|  | Techniques |  |  |  |  |  |  |
|  | NGS | Sanger | HRM | IHC | FISH | WGS | Total cost per cancer patient |
|  | PGM 316, 318 chip; MiSeq | ABI3500 (10/3/6/9 amplicons) | BRAF+NRAS | ALK+ROS1 | ALK+ROS1+RET |  |  |
| **NSCLC** |  |  |  |  |  |  |  |
| Test 1 | € 283.95 |  |  | € 203.77 |  |  | € 487.72 |
| Test 2 | € 283.95 |  |  |  | € 242.07 |  | € 526.01 |
| Test 3 |  | € 71.19 |  |  | € 242.07 |  | € 313.26 |
| **Melanoma** |  |  |  |  |  |  |  |
| Test 1 | € 283.95 |  |  |  |  |  | € 283.95 |
| Test 2 |  |  | € 74.56 |  |  |  | € 74.56 |
| Test 3 |  | € 57.68 |  |  |  |  | € 57.68 |
| **CRC** |  |  |  |  |  |  |  |
| Test 1 | € 283.95 |  |  |  |  |  | € 283.95 |
| Test 2 |  | € 63.47 |  |  |  |  | € 63.47 |
| **GIST** |  |  |  |  |  |  |  |
| Test 1 | € 283.95 |  |  |  |  |  | € 283.95 |
| Test 2 |  | € 69.26 |  |  |  |  | € 69.26 |
| **All** |  |  |  |  |  |  |  |
|  |  |  |  |  |  | € 4,738.05 | € 4,738.05 |

Test 1, 2 (and 3) show the descending order of frequency usage of technique (combinations).
For NSCLC, the techniques included in test 1 are concomitantly applied (100 %) and those incorporated in test algorithm 2 and 3 are sequential applied (< 100 %), only when the prior test is negative.
The total cost for FISH is based on 60 % frequency usage of ALK, ROS1 and RET (sequential testing; in 40 % of cases a mutation in EGFR or KRAS is observed, and consequently FISH is not performed) (26-27).
The genes tested with Sanger are EGFR, KRAS, BRAF, ERBB2 and MET (10 amplicons) for NSCLC; BRAF and NRAS (3 amplicons) for melanoma; KRAS, NRAS and BRAF (6 amplicons) for CRC; KIT, PDGFRA and BRAF (9 amplicons) for GIST.
Including non-small cell lung carcinoma (NSCLC), colorectal cancer (CRC) and gastrointestinal stromal tumor (GIST).
☐ Including next generation sequencing (NGS), Sanger sequencing (Sanger), High Resolution Melting (HRM), immunohistochemistry (IHC), Fluorescence In Situ Hybridization (FISH) and whole genome sequencing (WGS).

### Sensitivity analysis

The sensitivity analyses demonstrate the impact on the total cost per cancer patient of varying platform utilization (+ / – 15 % standard techniques; + 30 % WGS) and reducing consumable cost by 30 %. In Table 4 the base case total cost, and the range resulting from varying platform utilization and reducing consumable cost, are shown per frequently occurring technique as included in Table 3. In any case, the ranges show overall cost reductions: to illustrate, from € 284 (average 32 % platform utilization) to € 250 (17 % platform utilization; – 30 % consumable cost) and € 216 (47 % platform utilization; – 30 % consumable cost) for NGS gene panels; from € 204 (average 30 % platform utilization) to € 166 (15 % platform utilization; – 30 % consumable cost) and € 161 (45 % platform utilization; – 30 % consumable cost) for IHC (ALK, ROS1); from € 4738 (average 60 % platform utilization) to € 3403 (90 % platform utilization; – 30 % consumable cost) for WGS. However, varying platform utilization has little impact compared to reducing consumable cost, which seems to have a large impact.

**Table 4.** Sensitivity analysis.

| Table 4. Sensitivity analysis. |  |  |  |  |  |  |  |  |  |
| --- | --- | --- | --- | --- | --- | --- | --- | --- | --- |
|  | Techniques<sup>2,3</sup> |  |  |  |  |  |  |  |  |
|  | NGS<sup>2</sup> | Sanger |  |  |  | HRM | IHC | FISH | WGS |
|  | *PGM 316, 318 chip; MiSeq* | *ABI3500 (10 amplicons)* | *ABI3500 (3 amplicons)* | *ABI3500 (6 amplicons)* | *ABI3500 (9 amplicons)* | *BRAF+NRAS* | *ALK+ROS1* | *ALK+ROS1+RET* |  |
| Standard case perspective | € 283.95 | € 71.19 | € 57.68 | € 63.47 | € 69.26 | € 74.56 | € 203.77 | € 403.45 | € 4,738.05 |
| Range of practice<sup>2</sup> | € 250.11 - 216.01 | € 65.98 - 65.07 | € 56.52 - 55.62 | € 60.57 - 59.67 | € 64.63 - 63.72 | € 73.86 - 64.36 | € 166.06 - 161.06 | € 369.96 - 323.00 | € 3,403.47 |

<sup>2</sup> Currently most frequently used techniques in cancer types of focus.
<sup>3</sup> The average total cost of the cost calculation outcomes for the NGS platforms and different hotspot panels is used.
<sup>4</sup> The utilization of the platforms varied from 17 to 47 % for NGS (average 32 %), 39 to 69 % for Sanger (average 54 %), 13 to 43 % for HRM (average 28 %), 15 to 45 % for IHC (average 30 %), 9 to 39 % for FISH (average 24 %) and 60 % to 90 % for WGS. The cost of consumables were reduced by 30 % for all the techniques.
<sup>5</sup> Including next generation sequencing (NGS), Sanger sequencing (Sanger), High Resolution Melting (HRM), immunohistochemistry (IHC), Fluorescence In Situ Hybridization (FISH) and whole genome sequencing (WGS).

## Discussion

This micro-costing study provides detailed and comparable up to date costs of currently used diagnostic techniques and WGS in the context of predictive analysis for four cancer types. The total cost per cancer patient per technique varied considerably. For the vast majority of techniques, the operational costs (process of analysis costs such as consumables and personnel) accounted for over 90 % of the total per cancer patient technique costs (including any target gene, hotspot panel or protein expression subdivisions).

Strengths of the study are that the interpretation of each included cost item per cost category in the measurement plan was aligned extensively for all included techniques with those parties involved. Furthermore, a consistent and uniform method was used in performing process-based cost calculations of the application of the different techniques. Finally, these cost outcomes can be used for (comparative) value assessments on current and new diagnostic techniques.

In sensitivity analyses, the input parameters were changed (utilization platform percentage and cost of consumables). A large decrease in costs can be achieved when costs for consumables might be lower in the future. Even when platform utilization was reduced, the final total cost was lower for all techniques compared to the calculated standard case perspective cost. This shows that the cost component consumables is a more important cost driver than platform utilization.

Comparing our cost outcomes with those initially presented in the literature indicate that our costs for standard techniques are relatively lower. Roughly estimated, the extent of reduction was 18 % for single-gene techniques (€ 265 – € 309 (12) versus on average € 65 – € 405 (Sanger, HRM, IHC, FISH, Biocartis, Cobas, Pyro seq calculations)) and 58 % for small TGPs of 5 to (∼) 50 gene panels (€ 376 – € 968 (12, 15–16) versus on average € 284 (NGS gene panels calculations)). An explanation for the differences in costs is most probably the different costing methods used and reference year. Our calculated price for WGS (€ 4738) is higher compared to the price by Wetterstrand ($ 4484 (about € 3870)) (19). Unfortunately, due to a lack of insight in pricing characteristics of previous calculations, we are not able to indicate reasons for this difference. Total cost for all DNA sequencing techniques including WGS is likely to decrease as a result of continuously ongoing innovations and due to market forces, leading to a reduced price of consumables over time, which is the main cost driver (17–18).

It should be stressed that total costs presented per technique are directly related to the test, data analysis and reporting process, so the final cost price indicated in this analysis is not the cost of what a technique costs in its entirety (exclusion of cost obtaining biopsy material, DNA extraction, VAT and overhead). Other excluded costs are, for example, time spend on training, validation, quality assurance and innovation costs (16). Noteworthy, novel genetic biomarkers are continuously emerging. However, these costs of development and implementation of new techniques, or adaptation and validation of existing techniques, are also excluded. Understandably, total cost of techniques is higher when focusing on the entire trajectory. Nonetheless, these outcomes are a snapshot in time, that is, all necessities (platforms, consumables) are strongly in development, especially for DNA sequencing techniques, and are likely to decrease in cost price (17–19). If so, this can easily be changed in our cost tables to calculate new cost prices.

Other factors that should be taken into account when comparing current standard diagnostic techniques and WGS are turnaround time, sensitivity, specificity, diagnostic yield and quality of the sequencing results. Among other things, the frequency of testing (e.g. number of sequencing runs per week or multiple, sequential, testing), the turnaround time of a sequencing run, time for data analysis and interpretation of results effects the total turnaround time of a sample. The success rate of the sequencing analysis depends on the quantity and quality (biopsy size, tumor volume) of the tumor material, and the amount and quality of DNA extracted (24–25). Subsequently, it determines the number of biopsies to be collected (new material needed when it turns out not to be of sufficient quantity or quality). Rationally, the latter in turn has an impact on waiting time till the start of treatment.

Some limitations of the study need to be addressed. First, this study made an attempt to define realistic assumptions that define the likely cost of application of these techniques in an average lab practice in the Netherlands. Second, for the Dutch labs, the base case assumptions came forth based on a maximum of three labs per included technique, so not all 24 individual labs that helped in defining the most frequently used techniques. As for the HMF facility, the base case assumptions on WGS testing, assuming an average Dutch lab practice, were verified with in-house experts. Lastly, no data could be obtained for MassArray, which was initially identified as a frequently used technique in treatment of NSCLC and CRC (by one lab), and was therefore not included.

## Conclusion

This study provided a detailed overview of all costing aspects and cost prices of current and new diagnostic techniques in treatment of NSCLC, melanoma, CRC and GIST in the Netherlands. Costs varied between € 58 (Sanger, 3 amplicons) to € 4738 (paired tumor-normal WGS). Costs for commonly used techniques per cancer type varied between € 58 (Sanger) and € 284 (NGS) for melanoma, € 63 (Sanger) and € 284 (NGS) for CRC, € 69 (Sanger) and € 284 (NGS) for GIST, and technique combinations for NSCLC ranged from € 313 (Sanger and FISH) to € 526 (NGS and FISH). The cost of WGS is significantly higher compared to the cost of standard techniques, but it is expected to decrease over time. In terms of value, diagnostic yield is potentially larger with WGS. Though, the study exclusively compared the different techniques based on cost price and not based on their potential value.

Differences in value were not collected in this study, therefore this study can and should be used as starting point in comparing diagnostic modalities. Important to note is that additional factors with regard to value ought to be included to fully assess added benefits (both on monetary as well clinical aspects) of new diagnostic techniques. Future economic evaluations of diagnostic modalities should take into account this difference in value together with the detail costing to give a more comprehensive meaning to the comparison of diagnostic techniques used in cancer treatment. These evaluations support decision-making on implementation of WGS and other diagnostic modalities in routine clinical practice.

## Data Availability

Data presented in the tables is the complete set of data

## Funding sources

This work is part of the research program Personalized Medicine, which is financed by the Netherlands Organisation for Health Research and Development (ZonMw, project numbers 846001001 and 846001002). Other grant providers are the HMF, the Dutch Cancer Society (KWF Kankerbestrijding) and the Dutch health care insurance company Zilveren Kruis.

## Acknowledgements

The authors would like to thank Wim van Harten, Manuela Joore, Martijn Simons, Erik Koffijberg, Maarten IJzerman, Michiel van de Ven and Inge Eekhout from the Technology Assessment of Next Generation Sequencing in Personalized Oncology (TANGO) consortium, and Astrid Eijkelenboom, Arja ter Elst, Robert van der Geize, Winand Dinjens, Carel van Noesel, Clemens Prinses, Ernst-Jan Speel from the Predictive Analysis for Therapy (PATH) consortium. Furthermore, they would like to express gratitude to the HMF facility and the Dutch pathology laboratories who participated in this study.

## Supplementary tables

**Table S1.** Percentage frequency usage techniques per cancer type (PALGA, 2018).⍰

| Table S1. Percentage frequency usage techniques per cancer type (PALGA, 2018).<sup>1</sup> |  |  |  |  |  |  |  |  |  |  |
| --- | --- | --- | --- | --- | --- | --- | --- | --- | --- | --- |
|  | Techniques<sup>2</sup> |  |  |  |  |  |  |  |  |  |
|  | NGS | NGS + other technique | MassArray | HRM | Sanger | Pyro seq | Biocartis | Cobas | Other | Unknown |
| NSCLC<sup>3</sup> | 78 | 5 | 3 | 4 | 5 | 3 | 1 | 0 | 0 | 0 |
| Melanoma | 57 | 8 | 15 | 6 | 5 | 1 | 4 | 1 | 1 | 0 |
| CRC<sup>3</sup> | 80 | 2 | 2 | 4 | 7 | 2 | 3 | 0 | 0 | 0 |
| GIST<sup>3</sup> | 78 | 16 | 0 | 0 | 6 | 0 | 0 | 0 | 0 | 0 |

<sup>1</sup> Online only.
<sup>2</sup> Including non-small cell lung carcinoma (NSCLC), colorectal cancer (CRC) and gastrointestinal stromal tumor (GIST).
<sup>3</sup> Including next generation sequencing (NGS), High Resolution Melting (HRM), Sanger sequencing (Sanger) and pyrosequencing (Pyro seq).

**Table S2.**
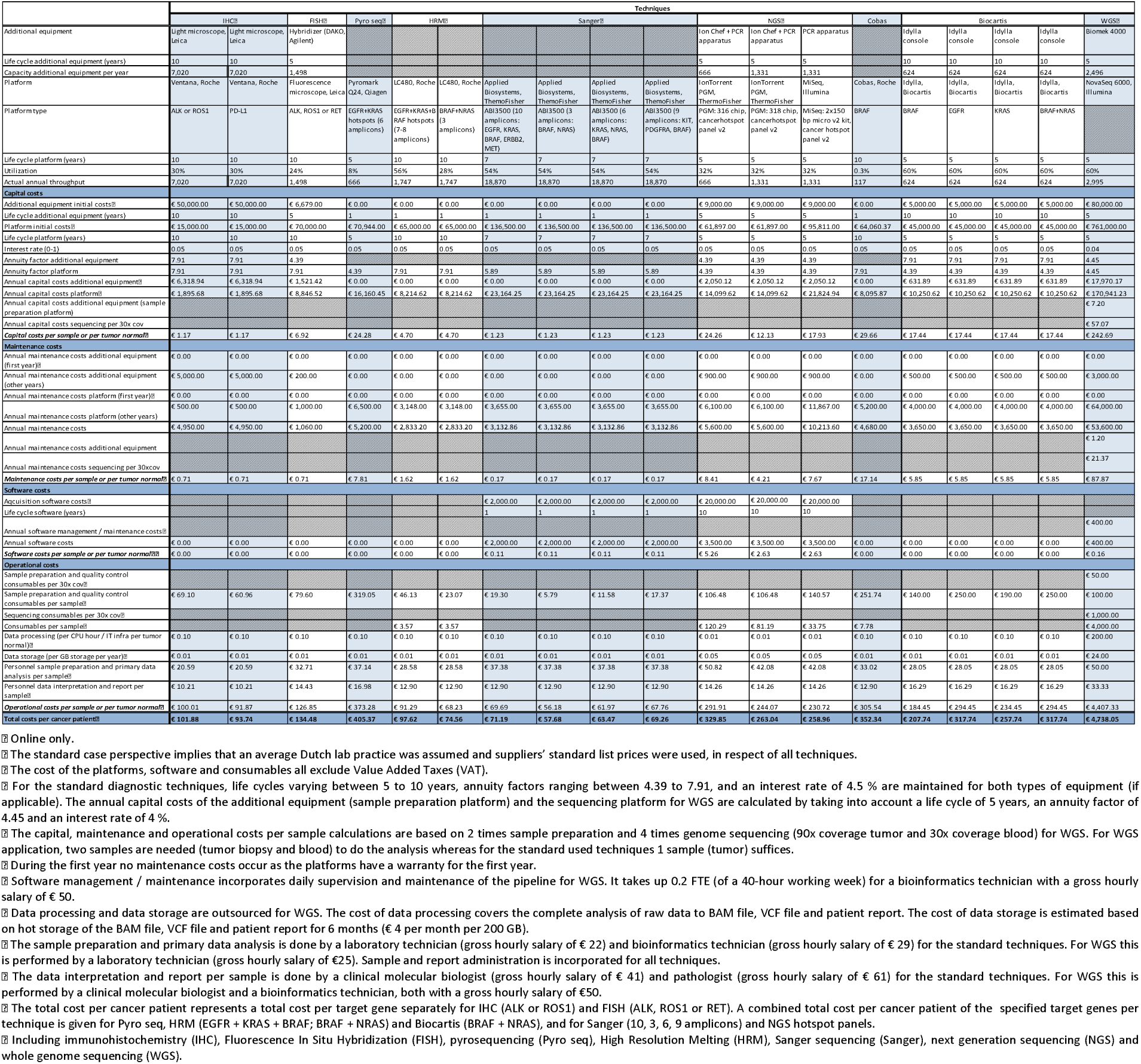
Process-based cost calculations of diagnostic applications based on the standard case perspective.⍰⍰

## Notes

### Competing Interest Statement

The authors have declared no competing interest.

### Author Declarations

All relevant ethical guidelines have been followed and any necessary IRB and/or ethics committee approvals have been obtained.

Any clinical trials involved have been registered with an ICMJE-approved registry such as ClinicalTrials.gov and the trial ID is included in the manuscript.

